# Performance of saliva specimens for the molecular detection of SARS-CoV-2 in the community setting: does sample collection method matter?

**DOI:** 10.1101/2020.12.01.20241349

**Authors:** Marta Fernández-González, Vanesa Agulló, Alba de la Rica, Ana Infante, Mar Carvajal, José Alberto García, Nieves Gonzalo-Jiménez, Claudio Cuartero, Montserrat Ruiz, Carlos de Gregorio, Manuel Sánchez, Mar Masiá, Félix Gutiérrez

## Abstract

**Background:** Data on the performance of saliva specimens for diagnosing COVID-19 in ambulatory patients are scarce and inconsistent. We assessed saliva-based specimens for detecting SARS-CoV-2 by RT-PCR in the community setting and compared three different collection methods.

**Method:** Prospective study conducted in three primary care centres. RT-PCR was performed in paired nasopharyngeal swabs (NPS) and saliva samples collected from outpatients with a broad clinical spectrum of illness. To assess differences in collection methods, saliva specimens were obtained in a different way in each of the participating centres: supervised collection (SVC), oropharyngeal washing (OPW) and self-collection (SC).

**Results:** NPS and saliva pairs of samples from 577 patients (median age 39 years, 44% men, 42% asymptomatic) were collected and tested, and 120 (20.8%) gave positive results. The overall agreement with NPS and kappa coefficients (KƘ) for SVC, OPW and SC were 95% (Ƙ=0.85), 93.4% (Ƙ=0.76), and 93.3% (Ƙ=0.76), respectively. The sensitivity (95% CI) of the saliva specimens varied from 86% (72.6-93.7) for SVC to 66.7% (50.4-80) for SC samples. The sensitivity was higher in samples with lower cycle threshold (Ct) values. The best performance of RT-PCR was observed for SVC, with sensitivity (95% CI) for Ct values ≤32 of 97% (82.5-99.8) in symptomatic, and 88.9% (50.7-99.4) in asymptomatic individuals.

**Conclusions:** Saliva is an acceptable specimen for the detection of SARS-CoV-2 in the community setting. Specimens collected under supervision perform comparably to NPS and can effectively identify individuals with higher risk of transmission in real life conditions.

## Introduction

SARS-CoV-2 infection is usually detected by real-time reverse transcriptase polymerase chain reaction (RT-PCR) on RNA extracted from upper respiratory tract specimens, typically nasopharyngeal swabs (NPS). However, collection of NPS is uncomfortable for the patient and may induce cough and sneezing, which may expose health care provider to infectious aerosols. Therefore, alternative sampling has been investigated, including easy-to-obtain specimens with the potential for patient self-collection such as saliva. Although initial studies investigating the use of saliva suggested that this specimen may be a good alternative sample to NPS, mixed results have been reported with sensitivities in the range of 30 to 100% [1–12].

Most of the studies that have evaluated SARS-CoV-2 detection in saliva were conducted in patients admitted to hospital with known COVID-19 infection and some of them were limited by the lack of simultaneous collection of NPS and saliva specimens and by the reduced composition of the cohorts, including mainly adults and symptomatic patients, all of which may limit the overall generalizability of the findings. Additionally, the procedure for collecting saliva specimens has varied substantially among the studies from enhanced collection under direction or supervision by the clinician [1-5] to unsupervised self-collection by the participants [10-12]. Variation in saliva sampling may be an explanation for the varying results of the published studies.

The use of saliva specimens in the ambulatory setting may be particularly appealing due to ease of collection and reduced equipment required, but data are scarce and inconsistent. While findings from a study on 45 patients support its potential for detecting SARS-CoV-2 from outpatients [5], a reduced sensitivity relative to NPS has been reported in a recent community-based cohort, raising concerns on the use of saliva samples in this setting [12,13]. Therefore, to clarify the role of saliva as an alternate specimen type for the detection of SARS-CoV-2 in the community setting, larger clinical studies are needed.

The present study aimed to evaluate the performance of saliva-based specimens for detecting SARS-CoV-2 by RT-PCR in a prospective study of adults and children with suspected COVID-19. In this investigation, we performed RT-PCR in paired NPS and saliva samples collected from outpatients with a broad clinical spectrum of illness, including asymptomatic cases, undergoing SARS-CoV-2 testing, and compared three different collection methods.

## Methods

### Setting and study subjects

This prospective, observational study was carried out at the Departments of Heath 17 and 20 of the province of Alicante, Spain. The study was approved by the Hospital General Universitario de Elche COVID-19 Institutional Advisory Board. Patients enrolled in the study were those presenting for SARS-CoV-2 testing as requested by their providers, at three primary care centres (PCC) facilities. Both symptomatic patients and asymptomatic subjects that had been exposed to SARS-CoV-2 were invited to participate in the investigation by providing saliva samples immediately preceding collection of the NPS. After obtaining written consent, demographic and clinical findings were recorded.

### Specimen collection and processing

Saliva specimens were collected into a 100 ml sterile empty container without transport medium. To assess differences in collection methods, saliva specimens were obtained in a different way in each of the participating PCC by random. In centre A, saliva specimens were obtained under the supervision of a healthcare worker (Supervised collection, SVC); in centre B, saliva specimens were obtained after oropharyngeal washing (OPW) with 2 ml of saline solution for 1-2 minutes prior to collection, and in centre C, saliva specimens were collected independently by the individual providing the sample following written instructions asking them to repeatedly spit up to a minimum of 1 ml saliva into the collection pot before the NPS (Self-collection, SC). In the SVC centre, subjects were instructed to clear saliva from back of the throat without coughing, pooling it their mouth for 1-2 minutes while touching with the tip of the tongue on both cheeks, and both gums, and then repeatedly spit a minimum of 1 ml saliva into the collection pot in the presence of a healthcare worker.

In the three PCC, nasopharyngeal samples were taken by qualified nurses following the same procedure. The flexible, mini-tip swab was passed through the patient’s nostril until the posterior nasopharynx was reached, left in place for several seconds to absorb secretions, then slowly removed while rotating. Swabs were placed in 3 ml of sterile transport media containing guanidine salt (Mole Bioscience, SUNGO Europe B.V., Amsterdam, Netherlands). Nasopharyngeal and saliva specimens were transported within 2 hours of sample collection to the clinical microbiology laboratory for molecular analysis by RT-PCR. NPS samples were analyzed immediately and saliva specimens frozen (−20 °C) and analyzed within two weeks after collection.

### SARS-CoV-2 detection

Nucleic acid extraction was performed using 300 µL of specimen (NPS or saliva) on Chemagic™ 360 Nucleic Acid Purification Instrument (PerkinElmer España SL, Madrid, Spain). Then, 10 uL of eluate was used for real-time RT-PCR assay targeting the E-gene (LightMix® Modular SARS-CoV (COVID19) E gene, TIB MOLBIOL, Berlin, Germany, distributed by Roche). Testing was performed according to the manufacturer’s guidelines on Cobas z 480 Analyzer (Roche, Basilea, Suiza). The number of cycles of amplification in RT-PCR (cycle threshold, Ct) value was assessed as a surrogate measure of the RNA concentration. Per the manufacturer, a Ct value of ≤40 by PCR is considered a positive result.

### Statistical methods

Continuous variables were expressed as median ± 25th and 75th percentiles (Q1, Q3), and categorical variables as percentages. Wilcoxon or Student’s t-test were used to compare two continuous variables and Kruskal Wallis anova for three or more, and the chi-square or Fisher’s exact test for categorical variables comparison.

The percent agreement (positive, negative and overall) (PPA, NPA and OPA) for saliva specimens obtained by SVC, OPW and SC was calculated using the results of the RT-RCR test in NPS as reference standard. Performance agreement was evaluated using kappa coefficients (κ). Positive results of either NPS or saliva specimens were considered true positives for calculations of sensitivity. Patients with an undetermined result in RT-PCR for NPS or saliva specimen were not considered for calculations of agreement. Performance of saliva specimens was also evaluated stratifying by age, Ct and presence of symptoms. Statistical analyses were performed with R version 4.0.3 (2020-10-10) software. Percent positive or negative agreement and kappa coefficients (κ) were calculated using the package caret [14]. For the graphical analysis, the ggplot2 package [15] was used.

We planned to include 133 patients per arm assuming a sensitivity of 95% and a confidence of 95%. With this sample size the study would have a statistical power of 80% to detect a 10% difference in sensitivity between the different collection methods.

## Results

### Patient characteristics and positivity rates

A total of 634 patients (103 children) were invited to take part in the study between 15th September and 29th October 2020. Fifty-four (44 children) were unable to provide saliva specimens and 3 (0.5%) specimens had insufficient sample for laboratory analysis. 577 pairs of samples (229 SVC, 140 OPW, 208 SC) were included in the analyses. Flow chart of the patients is depicted in Supplemental Figure S1. Demographic and clinical characteristics of the patients according to the collection method of saliva specimens are shown in Table 1. There were 120 (20.8%) positive results for SARS-CoV-2, 50 (21.8%) in the SVC, 28 (20%) in the OPW and 42 (20.2%) in the SC group. NPS and saliva samples from 2 (0.3%) and 9 (1.6%) patients, respectively, generated invalid transcription-mediated amplification results due to internal control failure.

**Table 1.** Demographic and clinical characteristics of the patients according to the collection method of saliva specimens.

|  | Overall cohort |  | Supervised collection (SVC) |  | Oropharyngeal washing (OPW) |  | Self-collection (SC) |  |
| --- | --- | --- | --- | --- | --- | --- | --- | --- |
|  | All patients | Positive SARS-Cov-2 RNA | SVC | Positive SARS-Cov-2 RNA | OPW | Positive SARS-Cov-2 RNA | SC | Positive SARS-Cov-2 RNA |
| No patients | 577 | 120 (20.8) | 229 | 50 (21.8) | 140 | 28 (20) | 208 | 42 (20.2) |
| Gender, male | 251 (43.5) | 57 (47.5) | 91 (39.7) | 22 (44) | 66 (47.1) | 13 (46.4) | 94 (45.2) | 22 (52.4) |
| Age (years), median (Q1-Q3) | 39 (24-51) | 42 (28-54) | 39 (21-48) | 42 (29-49) | 36 (23-51) | 41 (27-54) | 40 (27-54) | 41 (28-57) |
| 0-14 | 59 (10.2) | 5 (4.2) | 28 (12.2) | 3 (6) | 16 (11.4) | 1 (3.6) | 15 (7.2) | 1 (2.4) |
| 15-50 | 368 (63.8) | 79 (65.8) | 152 (66.4) | 35 (70) | 87 (62.1) | 18 (64.3) | 129 (62) | 26 (61.9) |
| 50-65 | 105 (18.2) | 26 (21.7) | 40 (17.5) | 10 (20) | 26 (18.6) | 8 (28.6) | 39 (18.8) | 8 (19) |
| >65 | 45 (7.8) | 10 (8.3) | 9 (3.9) | 2 (4) | 11 (7.9) | 1 (3.6) | 25 (12) | 7 (16.7) |
| Hypertension | 42 (7.3) | 13 (10.8) | 17 (7.4) | 3 (6) | 12 (8.6) | 2 (7.1) | 13 (6.2) | 8 (19) |
| Dyslipidemia | 37 (6.4) | 15 (12.5) | 17 (7.4) | 5 (10) | 11 (7.9) | 5 (19.2) | 9 (4.3) | 5 (11.9) |
| Diabetes | 19 (3.3) | 9 (7.5) | 10 (4.4) | 3 (6) | 4 (2.9) | 3 (10.7) | 5 (2.4) | 3 (7.1) |
| Cardiomyopathy | 17 (2.9) | 3 (2.5) | 9 (3.9) | 0 (0) | 7 (5) | 2 (7.1) | 1 (0.5) | 1 (2.4) |
| Obesity | 25 (4.3) | 10 (8.3) | 9 (3.9) | 2 (4) | 8 (5.7) | 3 (10.7) | 8 (3.8) | 5 (11.9) |
| Symptomatic | 336 (58.2) | 86 (71.7) | 145 (63.3) | 37 (74) | 68 (48.6) | 17 (60.7) | 123 (59.1) | 32 (76.2) |
| Days with symptoms, median (Q1-Q3) | 4 (3-6) | 5 (3-7) | 3 (2-5) | 4 (2-7) | 4 (3-5) | 4 (3-6) | 4 (3-6) | 5 (3-7) |
| **Sensitivity of saliva specimens (95% CI)** |  |  |  |  |  |  |  |  |
| Global | 76.7% (67.9-83.7) |  | 86% (72.6-93.7)* |  | 75% (54.8-88.6) |  | 66.7% (50.4-80) |  |
| Symptomatic | 77.9% (67.4-85.9) |  | 89.2% (73.6-96.5)** |  | 70.6% (44-88.6) |  | 68.8% (49.9-83.3) |  |
| Asymptomatic | 73.5% (55.3-86.5) |  | 76.9% (46-93.8) |  | 81.8% (47.8-96.8) |  | 60% (27.4-86.3) |  |
| Ct ≤ 25 | 96.7 % (87.5-99.4) |  | 100% (85.9-100) |  | 93.3% (66-99.7) |  | 93.3% (66-99.7) |  |
| Ct ≤ 30 | 91.6% (82.9-96.3) |  | 97.4% (84.9-99.9) |  | 81% (57.4-93.7) |  | 91.3% (70.5-98.5) |  |
| Ct > 30 | 43.2% (27.5-60.4) |  | 45.5% (18.1-75.4) |  | 57.1% (20.2-88.2) |  | 36.8% (17.2-61.4) |  |

Categorical variables are represented by number and (%); COPD, chronic obstructive pulmonary disease; Ct, cycle threshold of RT-PCR.
\*P = 0.028 for the comparison between supervised collection and self-collection.
\*\*P=0.035 for the comparison between supervised collection and self-collection.

### Sensitivity of the different specimens for SARS-CoV-2 detection and concordance between saliva specimens and NPS

Table 2 shows the qualitative positive results for SARS-Cov-2 RNA obtained from NPS and saliva specimens according to the collection method, and Figure 1 the concordance of positive test results between NPS and the different saliva specimen types. The sensitivity for NPS specimens was 95% (95% CI, 88.9-97.9). Among saliva specimens, SVC showed the best case detection rate (43 of 50 infected patients) with significantly higher sensitivity than SC samples (86% [95% CI, 72.6-93.7] versus 66.7% [50.4-80]; p=0.027). OPW detected 21 of 28 individuals (sensitivity, 75% [95% CI], 54.8-88.6) (Table 1). The greatest sensitivity was obtained by combining NPS sampling with saliva collected under supervision (sensitivity 97.5% [95% CI, 92.3-99.3]). Table 3 shows the agreement of the three different saliva specimens with NPS. The best agreement with NPS was found for the specimens of the SVC group with a kappa coefficient of 0.85. For the OPW and SC groups, the kappa coefficient was 0.76.

**Table 2.** Comparison of qualitative results from nasopharyngeal swabs and saliva specimens according the collection method.

| **Saliva** |  | **Supervised collection**<br>(n=229) |  |  | **Oropharyngeal washing**<br>(n=140) |  |  | **Self-collection**<br>(n=208) |  |  |
| --- | --- | --- | --- | --- | --- | --- | --- | --- | --- | --- |
|  |  | Pos | Neg | Und | Pos | Neg | Und | Pos | Neg | Und |
| **Nasopharyngeal swab** | Pos | 39 | 7 | 0 | 18 | 7 | 0 | 28 | 14 | 0 |
|  | Neg | 4 | 171 | 7 | 2 | 110 | 2 | 0 | 166 | 0 |
|  | Und | 0 | 1 |  | 1 | 0 |  | 0 | 0 |  |

Pos, positive; Neg, negative, Und, undetermined.

**Table 3.** Agreement of the different saliva specimens with the nasopharyngeal swabs.

| **Saliva type** | **Positive percent agreement (95% CI)** | **Negative percent agreement (95% CI)** | **Overall percent agreement (95% CI)** | **Performance Agreement (Kappa) (95% CI)** |
| --- | --- | --- | --- | --- |
| **Supervised collection** | 84.8%<br>(70.5-93.2) | 97.7%<br>(93.9-99.3) | 95%<br>(91-97.4) | 0.85 (0.76-0.93) |
| **Oropharyngeal washing** | 72%<br>(50.4-87.1) | 98.2%<br>(93.1-99.7) | 93.4%<br>(87.5-96.8) | 0.76 (0.61-0.91) |
| **Self-collection** | 66.7%<br>(50.4-80) | 100%<br>(97.2-100) | 93.3%<br>(88.7-96.1) | 0.76 (0.64-0.88) |

**Figure 1.**
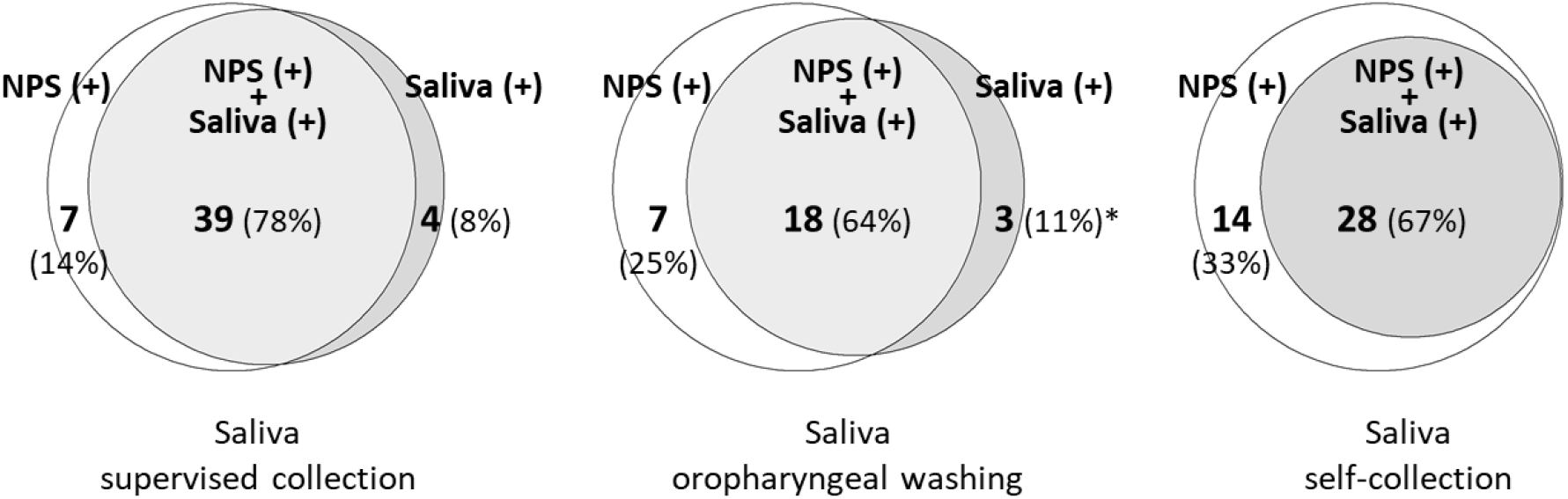
Positivity for SARS-CoV-2 RNA in nasopharyngeal swabs (NPS) and in the different saliva specimens. *One positive saliva, with NPS negative confirmed six days later.

### Performance of the different saliva specimens for SARS-CoV-2 detection by cycle threshold values and presence of symptoms

Median Ct values were significantly lower in NPS than in their paired saliva specimens (p=0.035), although in 19 (15.8%) patients the saliva showed a lower Ct than the corresponding NPS. Median Ct values in the three saliva specimen types were not significantly different from each another (p=0.962) (Figure 2). Supplemental Figure S2 displays the concordance for SARS-CoV-2 detection between paired NPS and saliva samples, and Ct values of the discordant positive results. Median (Q1-Q3) Ct values for NPS-positive-only or saliva-positive-only specimens were 33 (31-34) and 32 (29-33.5), respectively.

**Figure 2.**
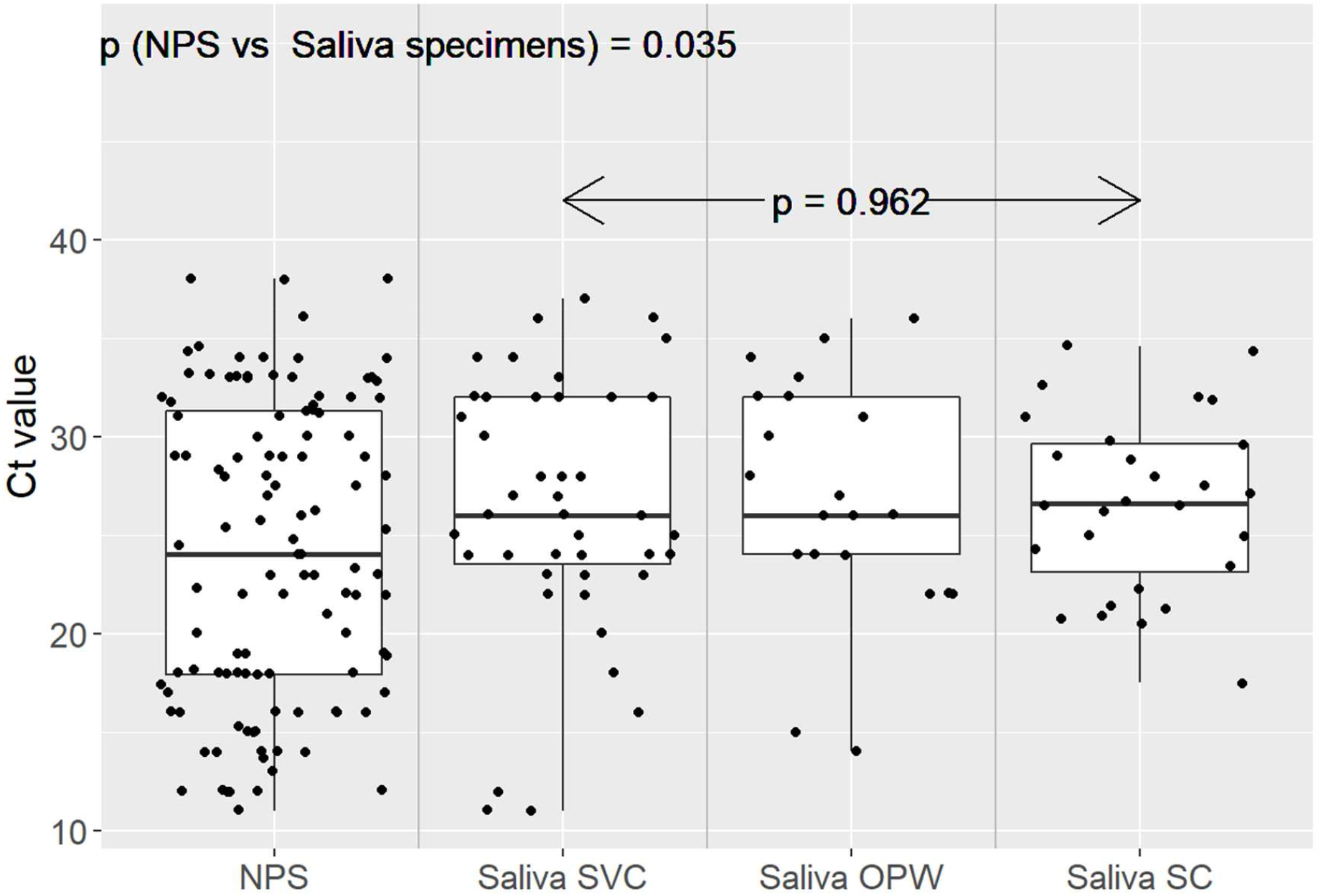
Matched Ct values for nasopharyngeal swabs and saliva specimens from all positive individuals collected. Each dot represents the Ct value (RT-PCR) for positive: nasopharyngeal swabs (NPS, n=113), saliva obtained under supervised collection (SVC, n=43), saliva obtained after oropharyngeal washing (OPW, n=21) and saliva obtained by self-collection (SC, n=28).

Sensitivity of the different specimens for SARS-CoV-2 detection according to Ct values is depicted in Figure 3. The sensitivity of the saliva specimens was higher in the samples with lower Ct values. For Ct values ≤ 25, median (95% CI) sensitivities of SVC, OPW and SC sampling reached 100% (85.9-100), 93.3% (66-99.7) and 93.3% (66-99.7), respectively, and decreased only minimally, to 97.4% (84.9-99.9), 81% (57.4-93.7) and 91.3% (70.5-98.5), respectively, for Ct ≤ 30. Supplemental Figure S3 shows the sensitivity by sample type according to Ct values and the presence of symptoms.

**Figure 3.**
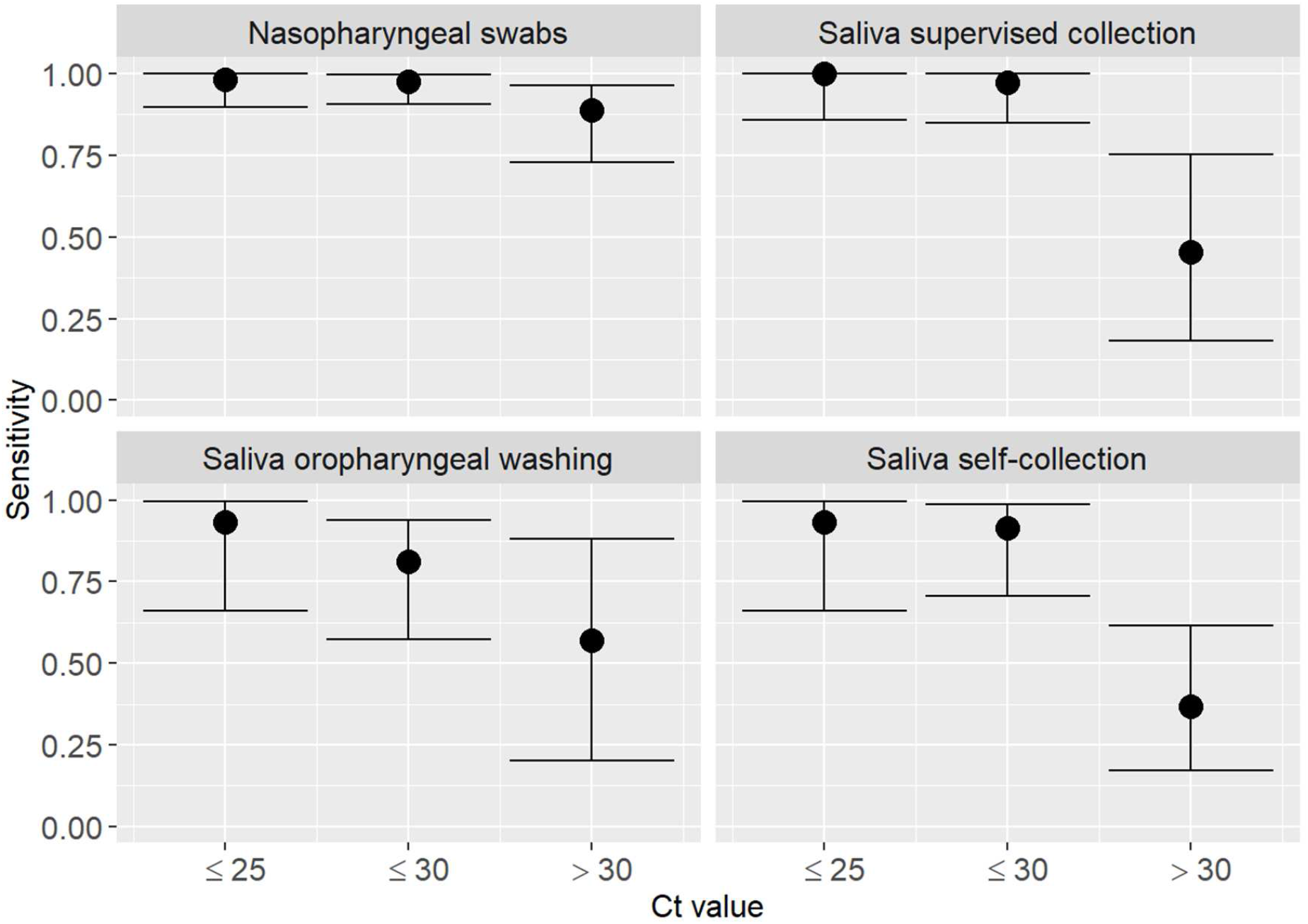
Sensitivity of the different specimens for SARS-CoV-2 detection according to Ct value.

There were no significant differences in the sensitivity between patients with and without active symptoms for the same Ct values across the different specimens. The best performance of RT-PCR was observed for NPS, closely followed by supervised collected saliva, with sensitivity for Ct values ≤ 32 of 95.8% (87.5-98.9) and 97% (82.5-99.8), respectively, in symptomatic individuals, and 95.7% (76-99.8) and 88.9% (50.7-99.4), respectively, in asymptomatic individuals.

## Discussion

We confirmed that saliva is an acceptable specimen for molecular detection of SARS-CoV-2 in the community setting and can effectively identify individuals with the highest risk of transmission in real life conditions. The study revealed that the collection method may be critical for improving sensitivity. Saliva specimens obtained under supervision outperforms self-collected samples and show higher sensitivity in symptomatic and asymptomatic patients.

As expected, the sensitivity of the saliva specimens increased in samples with low Ct values. Indeed, supervised collected specimens performed almost as well as nasopharyngeal samples with sensitivities well above 90% in patients with low Ct values, who are considered to have the greatest potential to spread the virus. For Ct values ≤ 25, the sensitivities of SVC, OPW and SC saliva specimens reached 100%, 93% and 93%, respectively, and they were 97%, 81% and 91% in patients with Ct values ≤ 30. Interestingly, in the participants whose saliva specimens tested negative and had positive results by NPS, the median Ct value was above 32, a threshold at which the risk for transmission is considered negligible [16]. Among NPS-only positive individuals we found only three cases of saliva collected under supervision who had Ct values ≤ 32. Therefore, the use of saliva specimens in general, and particularly when is obtained under supervision, allowed the detection of SARS-CoV-2 in the vast majority of the patients with significant risk of transmission.

In contrast to most of the previous studies, largely focused on inpatient populations, in the present investigation we included outpatients with a broad clinical spectrum of the illness, comprising children and asymptomatic cases. Like Williams et al. [11] we evaluated casual saliva specimens without previous fasting or enhancement techniques such as strong sniffing or coughing, used in other studies [4]. Prior investigations comparing different samples for molecular detection of SARS-CoV-2 in the community setting have reported lower sensitivity rates with saliva specimens than with NPS, ranging from 30% to 85% [11-13]. The reduced performance has been attributed to the milder symptoms in outpatients, with reduced viral load relative to more severe cases [17,18] and to differences in temporal dynamics in viral shedding in upper respiratory locations versus saliva [19,20] with lower viral loads in saliva samples [11]. We did not find prominent differences in the performance between those with or without active symptoms but, in line with other studies [8,21,22] we detected higher Ct values indicating lower viral loads in saliva specimens than in the corresponding NPS, which suggests differences in viral shedding between the two compartments. In addition, the significant differences in the performance of the specimen types evaluated in our study suggest that variation in saliva sampling may have contributed to the disparities in sensitivity observed in previous investigations.

The study has limitations. The investigation focused on comparing three specific methods for collection of saliva samples and was powered to detect rather large differences among groups. The sample size does not allow to draw conclusions on the performance in particular subgroups, including children and patients tested at different time point of illness. Noteworthy, a substantial proportion of the children recruited were unable to provide saliva, suggesting that this specimen might be less suitable for this group. In addition, we used a particular detection system (Cobas z 480 Analyzer), other platforms may have yielded different results. Strengths of the study are that it was population-based and carried out in real-life conditions, enrolling consecutive outpatients of all ages presenting for testing due to symptoms and asymptomatic people who had come into contact with confirmed cases.

In conclusion, our results indicate that the adequate collection of samples may be essential for the molecular diagnosis of COVID-19 when using saliva specimens. Saliva specimens obtained under supervision perform comparably to NPS and should be considered as a reliable sample for the diagnosis in the community setting in both symptomatic and asymptomatic individuals, particularly to detect individuals with significant risk of transmission. Although self-collected saliva would be the most advantageous way of sampling if mass testing were considered, these specimens had less sensitivity in our study. Further research is needed to determine whether other strategies of instruction, for example through videos or telematic means, can substitute for the direct supervision of a health professional.

## Data Availability

Data available within the article and its supplementary material

## Acknowledgements

*Members of the COVID19-Elx-Rapid Diagnostic Tests Study Group*

Félix Gutiérrez, Mar Masiá, Sergio Padilla, Guillermo Telenti, Lucia Guillen, María Andreo, Fernando Lidón, Vladimir Ospino, José López, Marta Fernández, Vanesa Agulló, Gabriel Estañ, Javier García, Cristina Martínez, Leticia Alonso, Joan Sanchís, Ángela Botella, Paula Mascarell, María Selene Falcón, Sandra Ruiz, José Carlos Asenjo, Carolina Ding, Mar Carvajal, Inmaculada Candela, Jorge Guijarro, Cristina la Moneda, Cristina Jara, Raquel Mora, Juan Manuel Quinto, Sergio Ros, Daniel Canal, Pascual Pérez, Carolina Garrido, Manuel Sánchez, Jaime Sastre, Carlos de Gregorio, Francisco Carrasco, Juan Navarro, Andrés Navarro, Nieves Gonzalo, Clara Pérez, Adoración Alcalá, José Luis Rincón, Montserrat Ruiz, Juan Antonio Gutiérrez.

