## Supplemental Figure S1, S2 and S3 for "Performance of saliva specimens for the molecular detection of SARS-CoV-2 in the community setting: does sample collection method matter?"

### Supplemental Data

Figure S1. Flow chart of the study.

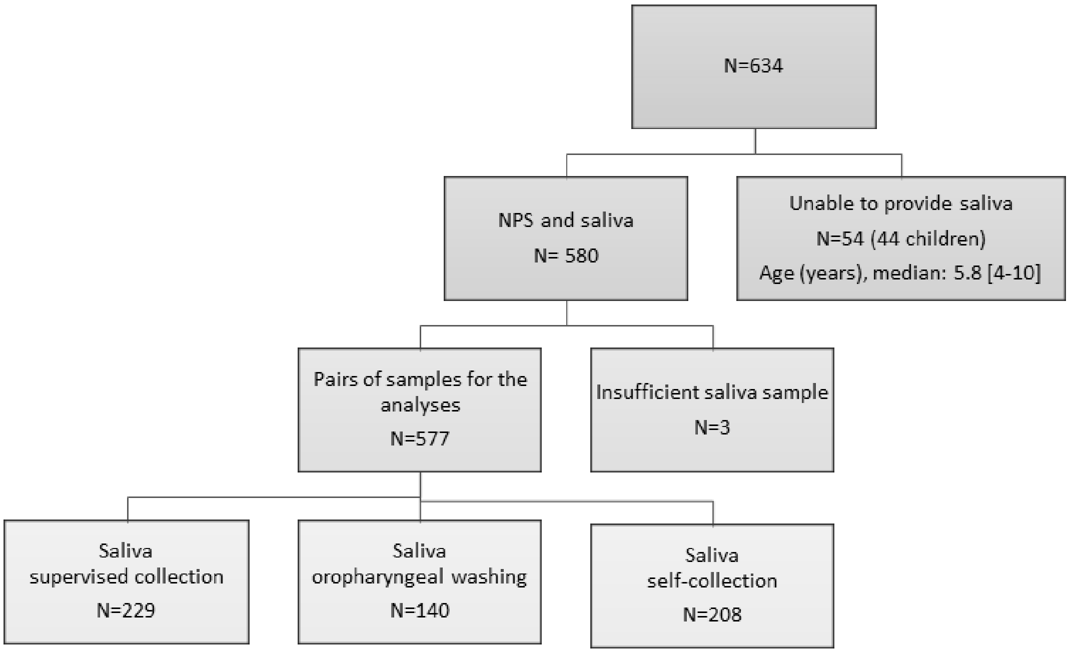


Figure S2. Comparison of SARS-CoV-2 detection by sample type and Ct value of discordant specimens.

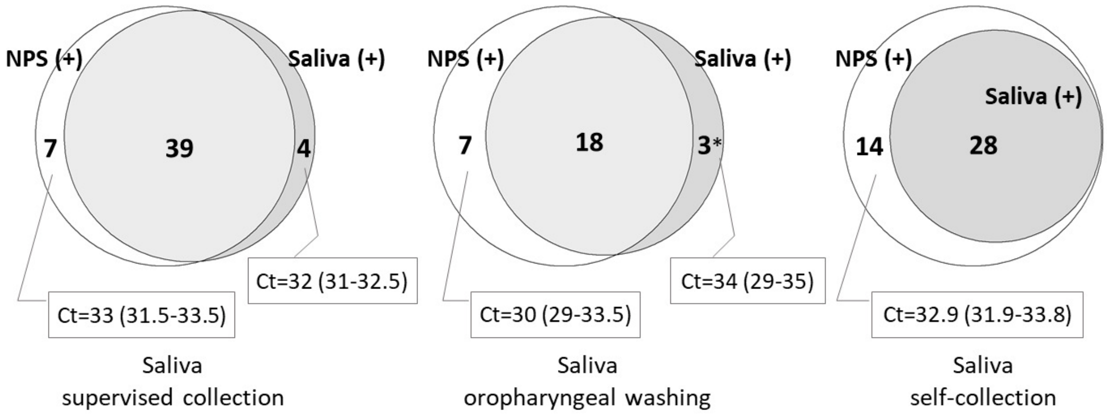


*One positive saliva, with NPS negative confirmed six days later.

Figure S3. Sensitivity of the different specimens for SARS-CoV-2 detection according to Ct value and presence of symptoms.

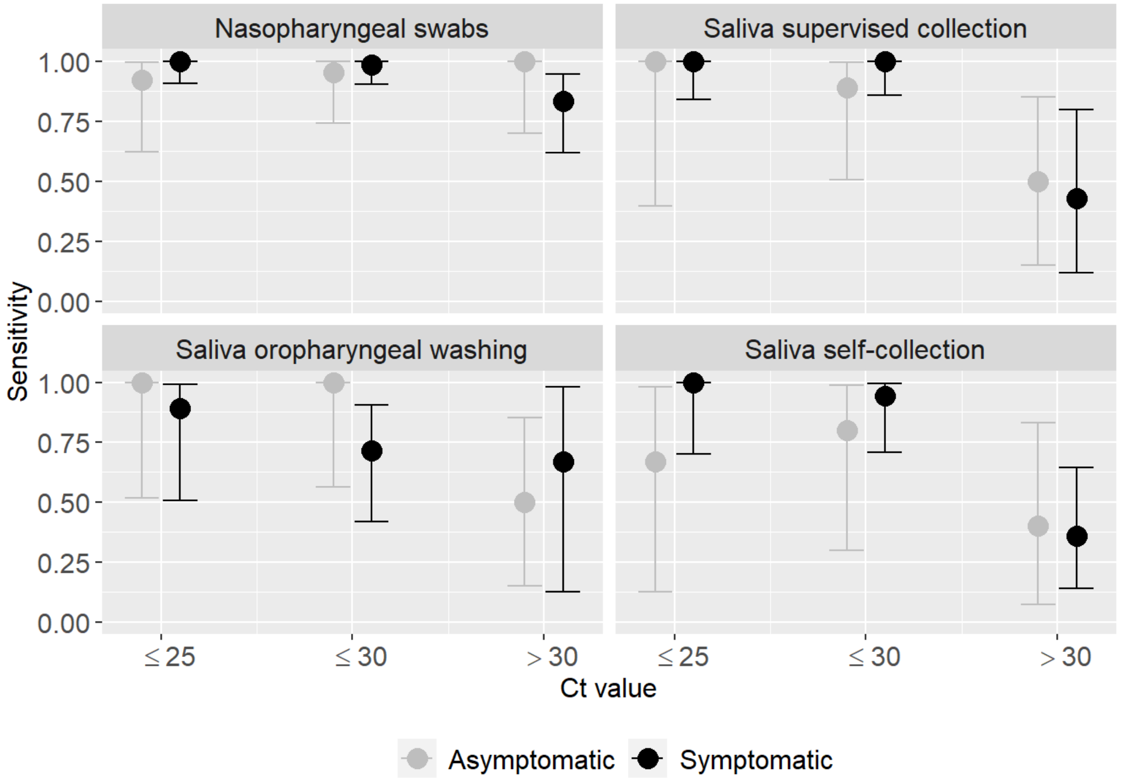
